# Measuring the missing: greater racial and ethnic disparities in COVID-19 burden after accounting for missing race/ethnicity data

**DOI:** 10.1101/2020.09.30.20203315

**Authors:** Katie Labgold, Sarah Hamid, Sarita Shah, Neel R. Gandhi, Allison Chamberlain, Fazle Khan, Shamimul Khan, Sasha Smith, Steve Williams, Timothy L. Lash, Lindsay J. Collin

## Abstract

Black, Hispanic, and Indigenous persons in the United States have an increased risk of SARS-CoV-2 infection and death from COVID-19, due to persistent social inequities. The magnitude of the disparity is unclear, however, because race/ethnicity information is often missing in surveillance data. In this study, we quantified the burden of SARS-CoV-2 infection, hospitalization, and case fatality rates in an urban county by racial/ethnic group using combined race/ethnicity imputation and quantitative bias-adjustment for misclassification. After bias-adjustment, the magnitude of the absolute racial/ethnic disparity, measured as the difference in infection rates between classified Black and Hispanic persons compared to classified White persons, increased 1.3-fold and 1.6-fold respectively. These results highlight that complete case analyses may underestimate absolute disparities in infection rates. Collecting race/ethnicity information at time of testing is optimal. However, when data are missing, combined imputation and bias-adjustment improves estimates of the racial/ethnic disparities in the COVID-19 burden.

## Introduction

In the United States, early surveillance reports highlight that persons of Hispanic, Black, and American Indian/Alaskan Native race and ethnicity are disproportionately affected by the COVID-19 pandemic.^1^ These disparities arise from historical and contemporary social and health inequities that result from systemic racism.^2–4^ Racial capitalism in particular produces structurally unequal exposure to (and protection from) SARS-CoV-2 infection in key places of transmission (*e*.*g*. workplace).^3^

The role of systemic racism in the pandemic motivates the need for accurate surveillance of racial/ethnic disparities in SARS-CoV-2 infection and death. However, there are challenges in estimating COVID-19 racial/ethnic disparities.^5,6^ Although reports highlight the unequal burden across racial/ethnic groups, the magnitude of disparities is uncertain because of the large proportion of missing race/ethnicity information in surveillance data. In recent reports, race/ethnicity information was missing in 56% of confirmed infections nationally and in 36% in Georgia.^7,8^ Current surveillance estimates are reported as complete case analyses, which exclude cases with missing race/ethnicity.^1,5,8,9^ Complete case analyses will bias racial/ethnic disparity estimates if race/ethnicity information is not missing completely at random.^10^

The Department of Health and Human Services issued COVID-19 reporting guidelines in June requiring all labs to report race/ethnicity beginning August 2020.^11^ These guidelines seek to address missing data moving forward, but fail to address missing information for case-patients identified before August.

Collecting race/ethnicity information at time of testing is optimal, especially in surveillance of racial/ethnic health disparities. Until this becomes routine, imputation of missing race/ethnicity combined with quantitative bias-adjustment to account for misclassification of the imputed race/ethnicity can improve estimates of the COVID-19 burden among racial/ethnic groups when race/ethnicity data are missing.^12^ In this study, we calculate SARS-CoV-2 infection, hospitalizations, and case fatality rates by race/ethnicity group and report the absolute racial/ethnic disparities in SARS-CoV-2 infection rates in Fulton County, Georgia after accounting for missing race/ethnicity information.

## Methods

Fulton County, Georgia, includes the city of Atlanta and residents identify as Black (44%), White (40%), Hispanic (7%), Asian (7%), and other races/ethnicities (2%).^13^ Between 29 February 2020 and 18 Aug 2020, 19,637 cases of SARS-CoV-2 infection were reported among Fulton County residents. Case reports included the patients’ residential address, full name, race/ethnicity, hospitalization (yes/no/unknown), and death (yes/no/unknown). Fulton County Board of Health staff geocoded case-patients’ address to census block groups. For this analysis, we categorized reported race/ethnicity as Black, Hispanic, Asian, White or Other.

We accounted for missing race/ethnicity information using a three-step approach: 1) <u>imputation</u> of race/ethnicity for all case-patients, 2) <u>validation</u> of the race/ethnicity imputation by calculating the accuracy of imputation among case-patients with reported race/ethnicity information, and 3) <u>bias-adjustment</u> of race/ethnicity estimates to account for misclassification of imputation among case-patients missing reported race/ethnicity information. Hereafter, we refer to race/ethnicity as reported when provided in case-patient records, *imputed* when referring to the imputed case-patient race/ethnicity, and *classified* when referring to the combined reported and imputed race/ethnicity after bias-adjustment.

First, for all case-patients we predicted their racial/ethnic group using the Bayesian Improved Surname Geocoding method.^14^ This method estimates the probability of a person being classified as Black, Hispanic, Asian, White or Other race/ethnic group based on the case-patient’s surname and residential census block group, and the population distribution of race/ethnicity for census block groups and surnames. Imputation was performed using the R package “wru,” which includes the 2010 surname census list with corresponding race/ethnicity distribution. Geographic distribution of race/ethnicity came from the 2018 5-year American Community Survey.^15,16^ For the 546 (2.8%) case-patients who could not be geocoded, race/ethnicity was imputed using surname only.

Second, we validated the race/ethnicity imputation among case-patients whose race/ethnicity was available in the dataset (n=12,222, 64%). Predictive values (PV) were calculated for each imputed race/ethnic group. The PV is the probability that a person’s reported race/ethnicity group classification was correctly imputed.^12^

Third, we used the PV values as bias parameters to quantitatively adjust for the expected misclassification of the imputed race/ethnicity groups. We assigned each race/ethnicity group PV from the validation to a Dirichlet distribution (**Table 1**). We then reclassified the imputed race/ethnicity probabilistically (100,000 iterations).^12^ The quantitative bias-adjustment mathematically accounts for inaccurate assignment of case-patients to a race/ethnicity group by the Bayesian Improved Surname Geocoding method. Sampling error was incorporated into the estimates using bootstrap approximation from a standard normal distribution.^12^

**Table 1:** Predictive values (PV) and 95% confidence intervals (CI) of the imputation by race/ethnicity based on residence and surname compared with reported race/ethnic group in the State Electronic Notifiable Disease Surveillance System

|  |  | Imputed Race/Ethnicity |  |  |  |  |
| --- | --- | --- | --- | --- | --- | --- |
|  |  | Black | Hispanic | Asian | White | Other |
| Reported Race/Ethnicity | Black | 5118 | 68 | 13 | 1754 | 11 |
|  | Hispanic | 77 | 1288 | 16 | 230 | 6 |
|  | Asian | 16 | 15 | 145 | 80 | 4 |
|  | White | 192 | 103 | 28 | 2827 | 2 |
|  | Other | 132 | 68 | 12 | 302 | 1 |
|  | Total | 5,535 | 1,543 | 214 | 5,193 | 24 |
| PV %<br>(95% CI) |  | 93%<br>(92%, 93%) | 84%<br>(82%, 85%) | 69%<br>(61%, 74%) | 55%<br>(53%, 56%) | 3.8%<br>(0.1%, 15%) |

For both the complete case and bias-adjusted analyses, we calculated the SARS-CoV-2 infection rates (per 1,000 persons), hospitalization proportions (hospitalized cases/reported cases), and case fatality rates (deaths/reported cases) by race/ethnicity group. We reported 95% confidence intervals (CI) for the complete case analysis and medians with 95% simulation intervals (SI) for the bias-adjusted estimates. We evaluated how accounting for missing race/ethnicity information impacts measures of racial/ethnic disparities by calculating the differences in SARS-CoV-2 infection rates in each race/ethnicity group compared with persons of White race/ethnicity, among case-patients with reported race/ethnicity information, and among all case-patients after bias-adjustment. All analyses used R v3.6 (Vienna, Austria). The Georgia Department of Health determined this activity to be consistent with public health surveillance, so does not require informed consent or IRB approval.

## Results

Among the 19,637 cases reported in Fulton County from 29 February to 19 August 2020, 7,145 (36%) were missing race/ethnicity information in the case report. Data were more complete among the 1,840 hospitalized case-patients, where only 14 (3.5%) were missing race/ethnicity information. All deceased case-patients (n=456) had complete information on race/ethnicity.

Comparison of reported versus imputed race/ethnicity group showed that the algorithm’s imputation accuracy varied by imputed race/ethnicity group (**Table 1**). Of the 5,535 persons who were imputed as Black race/ethnicity, 93% (95%CI: 92%, 93%) were reported as Black in case reports (n=5,118). Among persons imputed as Hispanic ethnicity, 84% (95%CI: 82%, 85%) were reported as Hispanic. The algorithm was less accurate for case-patients with race/ethnicity imputed as Asian (PV=69%, 95%CI: 61%, 74%) and as White (PV=55%, 95%CI: 53%, 56%). The PV estimates for racial/ethnic groups changed over time, likely due to changes in the prevalence of demographic groups affected by the pandemic over time (**Supplemental Table 1**).

In both the complete case and bias-adjusted analyses, the SARS-CoV-2 infection rates were highest among those classified as Other, followed by Hispanic, Black, White, and Asian (**Table 2a and 2b**). Imputation and bias-adjustment yielded higher estimates of infection rates than complete case analysis because more case-patients were included in the numerator. Estimated infection rates increased 1.8-fold for persons classified as Asian, 1.7-fold for White, 1.7-fold for Hispanic, 1.6-fold for Other, and 1.5-fold for Black. Hospitalization proportions and case fatality rates decreased across all race/ethnicity groups with imputation and bias-adjustment compared with the complete case analyses, because more cases were included in the denominator. In both the complete case and bias-adjusted analyses, case-patients who were classified as Black race/ethnicity had the highest hospitalization proportions (complete case: 17%, 95%CI: 16%, 18%; bias-adjusted: 12%, 95%SI: 11%, 12%) and case fatality rates (complete case: 4.6%, 95%CI: 4.1%, 5.1%; bias-adjusted: 3.1%, 95%SI: 2.8%, 3.4%).

**Table 2a:** Complete case estimates of SARS-CoV-2 infection rates, hospitalization proportions, and case fatality rates by race/ethnic group among 12,222 cases reported to Fulton County Board of Health, 29 February – 18 Aug 2020.

| Race/<br>Ethnicity | Total<br>infections | Hospitalized | Died | At Risk* | Infection rate per<br>1,000<br>(95%CI) | Hospitalized<br>percentage<br>(95%CI) | Case Fatality Rate as<br>a percentage<br>(95%CI) |
| --- | --- | --- | --- | --- | --- | --- | --- |
| Asian | 260 | 25 | 5 | 69987 | 3.7 (3.3, 4.2) | 9.6 (6.2, 14) | 1.9 (0.4, 3.8) |
| Hispanic | 1617 | 214 | 15 | 74328 | 22 (21, 23) | 13 (12, 15) | 0.9 (0.5, 1.4) |
| Black | 6964 | 1195 | 320 | 445992 | 16 (15, 16) | 17 (16, 18) | 4.6 (4.1, 5.1) |
| White | 3152 | 312 | 112 | 406755 | 7.7 (7.4, 8.0) | 9.9 (8.9, 11) | 3.6 (2.9, 4.2) |
| Other | 515 | 30 | 4 | 6056 | 85 (78, 92) | 5.8 (3.9, 8.0) | 0.8 (0.2, 1.6) |

**Table 2b:** Bias-adjusted estimates of SARS-COoV-2 infection rates, hospitalization proportions, and case fatality rates including 7,415 cases with imputed race/ethnicity, among 19,637 cases reported to Fulton County Board of Health before 18 Aug 2020.

| Race/<br>Ethnicity | Total infections<br>(95%SI) | Hospitalized | Died | At Risk* | Infection rate<br>per 1,000<br>(95%SI) | Hospitalized<br>percentage<br>(95%SI) | Case Fatality Rate<br>as a percentage<br>(95%SI) |
| --- | --- | --- | --- | --- | --- | --- | --- |
| Asian | 456 (439, 474) | 25 | 5 | 69987 | 6.5 (5.9, 7.2) | 5.5 (3.4, 7.6) | 1.1 (0.1, 2.1) |
| Hispanic | 2,691 (2,661, 2721) | 214 | 15 | 74328 | 36 (35, 38) | 7.9 (6.9, 9.0) | 0.6 (0.3, 0.8) |
| Black | 10,838 (10,327, 10,428) | 1195 | 320 | 445992 | 23 (23, 24) | 12 (11, 12) | 3.1 (2.8, 3.4) |
| White | 5,303 (5,250, 5,356) | 312 | 112 | 406755 | 13 (13, 13) | 2.1 (1.7, 2.5) | 2.1 (1.7, 2.5) |
| Other | 837 (810, 865) | 30 | 4 | 6056 | 138 (128, 148) | 0.5 (0.0, 0.9) | 0.5 (0.0, 0.9) |

\*American Community Survey 5-year 2018 estimates

The magnitude of the absolute disparity—difference in SARS-CoV-2 infection rates for case-patients classified in each race/ethnicity group compared with case-patients classified White— increased in the bias-adjusted analysis relative to the complete case analysis for nearly all race/ethnicity groups (**Table 3**). When comparing bias-adjusted with complete case results, the absolute disparity in infection rates increased 1.3-fold among classified Black and 1.6-fold among classified Hispanic race/ethnicity groups in reference to case-patients classified as White.

**Table 3:** Relative difference (RD) of SARS-CoV-2 infection rates among minority groups compared with non-Hispanic White persons among cases with complete information and after accounting for missing race/ethnicity among 4004 SARS-CoV-2 infected persons reported to Fulton County before 20 May 2020.

| Race/<br>Ethnicity | Complete Case |  | Bias-Adjusted |  | Relative<br>change in<br>magnitude of<br>disparity |
| --- | --- | --- | --- | --- | --- |
|  | Infection rate per<br>1,000<br>(95%CI)* | RD per 1,000<br>(95%CI) | Infection rate per<br>1,000<br>(95%SI) | RD per 1,000<br>(95%SI) |  |
| Asian | 3.7 (3.3, 4.2) | −4.0 (−4.6, −3.5) | 6.5 (5.9, 7.2) | −6.5 (−6.8, −6.2) | 0.6 |
| Hispanic | 22 (21, 23) | 14 (13, 15) | 36 (35, 38) | 23 (23, 23) | 1.6 |
| Black | 16 (15, 16) | 7.9 (7.4, 8.3) | 23 (23, 24) | 10 (10, 11) | 1.3 |
| White | 7.7 (7.4, 7.8) | Reference | 13 (13, 13) | Reference |  |
| Other | 85 (78, 92) | 77 (70, 84) | 138 (128, 148) | 125 (121, 130) | 1.6 |

## Discussion

In this study, accounting for missing race/ethnicity information revealed greater differences in SARS-CoV-2 infection rates comparing most racial/ethnic groups with case-patients of White race. These results suggest that national estimates, which exclude case-patients with missing race/ethnicity information, may underestimate the magnitude of absolute racial/ethnic disparities in COVID-19 morbidity and mortality.^6,8^

Our results underscore the need for imputation combined with bias-adjustment. In our study population, the PV estimates indicated that imputation alone overestimated infections among case-patients classified as White and underestimated infections among case-patients classified as Black. Therefore, imputation alone would have been insufficient.

Both the complete case analysis and the bias-adjusted estimates demonstrate important absolute racial/ethnic disparities in the infection rates. The bias-adjusted estimates do not change our understanding of the direction of racial/ethnic disparities in the COVID-19 pandemic; however, the magnitude of racial/ethnic disparities changed meaningfully after bias-adjustment. In contrast, the hospitalization proportion and case fatality rate decreased across all classified race/ethnicity groups after accounting for missing race/ethnicity information because few hospitalized or deceased case-patients were missing race/ethnicity information. These results highlight the need for more complete reporting so that health equity and racial justice efforts aimed at addressing these disparities operate on the most accurate data possible.

The imputation of race/ethnicity has limitations. The Bayesian Improved Surname Geocoding algorithm limits the racial/ethnic groups that can be imputed to Black, Hispanic, Asian, White, or Other. The reliance on categories of ‘other’ is problematic for identifying and addressing disparities in other racial/ethnic populations (*e*.*g*. indigenous populations). Future studies should explore how accounting for missing race/ethnicity impacts other disease burden measures.

Our findings emphasize the importance of collecting complete race/ethnicity data at the time of testing, for the current pandemic and future outbreaks. When data are missing, Bayesian Improved Surname Geocoding combined with quantitative bias-adjustment provides better estimates of the racial/ethnic disparities in SARS-CoV-2 infection rates, hospitalization proportions, and case fatality rates.

## Data Availability

Due to patient confidentiality, data are only available upon request from the Fulton County Board of Health and with IRB approval from the Georgia Department of Public Health. Example code used to perform the imputation and bias-adjustment is available on GitHub (https://github.com/lcolli5/Adaptive-Validation).

## Appendix

**Supplemental Table 1:**
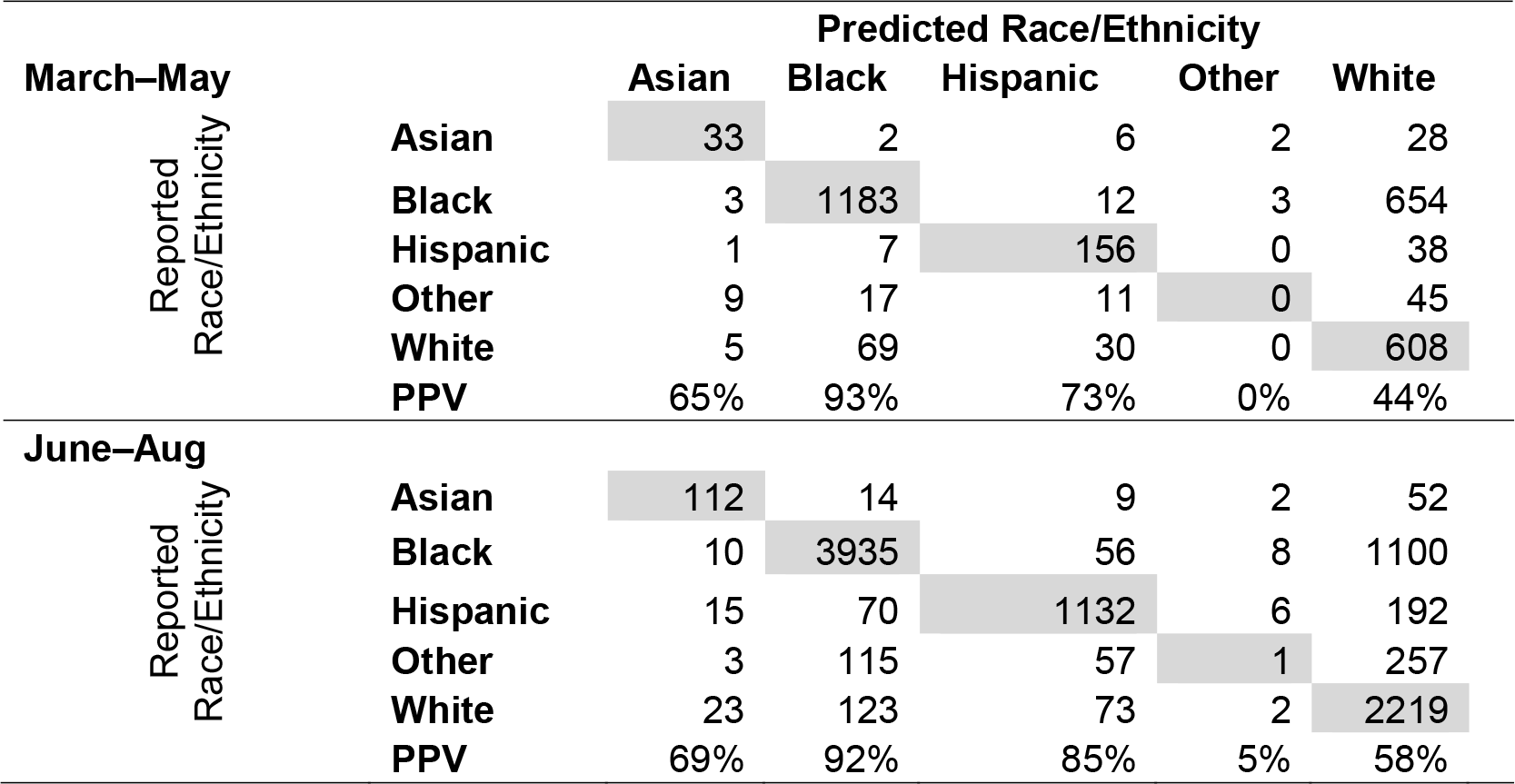
Positive predictive value (PPV) of the imputation by race/ethnicity based on residence and surname compared with the reported race/ethnic group in COVID-19 case report stratified by months (March through May and June through August) of diagnosis

